# Tongue swab Xpert MTB/RIF Ultra testing for tuberculosis in adolescents: a cross-sectional study of diagnostic accuracy and acceptability

**DOI:** 10.64898/2026.04.17.26351119

**Authors:** Emily Lai-Ho MacLean, Ma Thuy Thu, Chuong Huynh Long, Minh Huynh Khanh, Graeme Hoddinott, Pham Ngoc Yen, Tiep Trung Hua, Nguyen Thu-Anh, Greg J. Fox, Nguyen Thuy Ngoc

## Abstract

**Introduction:** Improved diagnostics are needed for people at risk of tuberculosis, especially adolescents. Tongue swab (TS) molecular testing has emerged as a promising strategy for tuberculosis diagnosis. We evaluated diagnostic accuracy and acceptability of Xpert MTB/RIF Ultra (Xpert) using TS samples for tuberculosis detection among adolescents.

**Methods:** We conducted a cross-sectional diagnostic accuracy study with consecutive recruitment in Vietnam. Adolescents aged 10-19 who were recommended to undergo investigation for tuberculosis and had not received tuberculosis treatment in the past years were eligible. Participants provided TS and sputum samples and completed a structured survey regarding sampling experiences. TS was tested on Xpert, with sputum tested on Xpert and liquid culture. We utilised a composite reference standard of a positive result on sputum Xpert or sputum culture to define disease status. Sensitivity, specificity, and diagnostic yield were calculated for TS Xpert.

**Results:** From July to December 2025, we enrolled 225 adolescents from Can Tho and An Giang provinces in southern Vietnam. Fewer than half (96/225, 43%) the participants exhibited a tuberculosis -like symptom, and the majority (157/225, 70%) were close contacts of a person recently diagnosed with tuberculosis. TS were collected from all adolescents, while 116 (52%) could provide mucopurulent sputum. Tuberculosis prevalence was relatively low (12/225, 5.3%). TS Xpert sensitivity (90% CI) and specificity (90% CI) were 58.3% (35.6, 78.0) and 99.5% (97.9, 99.9), respectively. Diagnostic yield among all diagnosed was 58.3% (7/12). TS sampling was highly acceptable to adolescents; the short time and simplicity of collecting TS were considered favourably.

**Conclusions:** The sensitivity and diagnostic yield of TS Xpert was relatively low among adolescents recommended for tuberculosis investigation, which includes asymptomatic individuals who may not provide high quality sputum. Specificity was excellent, and everyone could provide a TS. TS’s high acceptability indicates it remains a promising sample for diagnostic algorithms.

## Introduction

With over 10 million incident cases each year, but only eight million cases notified to national TB programmes (NTPs) (1), tuberculosis persists as a serious global health challenge.

Adolescents, defined by the World Health Organization (WHO) as people aged 10 to 19 years (2), are estimated to experience approximately 17% of new tuberculosis cases globally (3). WHO’s child and adolescent ‘TB Roadmap’ policy document calls for adolescent-specific TB strategies and recognises that their age-specific needs must be met to improve health outcomes (4). However, a lack of data may prevent countries from implementing approaches that are tailored to adolescents’ physical and social needs.

The importance of including children and adolescents in evaluating new diagnostic tools and approaches was highlighted in a recent consensus document (5). In particular, the document emphasises that tests utilising a pathogen-based measurand should be prioritised for evaluation in young people, as the target is consistent across ages and demographics. This rationale extends to the need to investigate alternative sample types, like tongue swabs (TS) with widely-available assays, such as Xpert MTB/RIF Ultra (Cepheid, USA) (Xpert) (6).

Promisingly, WHO has recently recommended the use of TS for tuberculosis testing on low complexity automated nucleic acid amplification tests, including Xpert, when respiratory samples, like sputum, cannot be obtained (7). The recommendation is applicable to adults and adolescents, although data informing the recommendation was mainly from adults with presumptive tuberculosis disease. A systematic review of molecular testing for tuberculosis using oral swabs included 20 studies, though only six recruited participants in the adolescent age range (8). Since the review’s publication, TS molecular testing has proven to be most accurate, and primary data is accumulating quickly, as evidenced by WHO’s new policy recommendation. However, an evidence gap persists regarding diagnostic accuracy of TS molecular testing in the adolescent age group specifically. To our knowledge, no published work has focussed specifically on TS molecular tuberculosis testing in an adolescent population. This means that adolescents may be having adult recommendations applied to them, without knowing if they are appropriate or fit-for-purpose (9).

Therefore, we aimed to estimate the diagnostic accuracy and yield of TS on Xpert MTB/RIF Ultra for detecting TB among adolescents aged 10 to 19 years in Vietnam. Additionally, we aimed to investigate adolescents’ experiences and acceptability of this testing scheme.

## Methods

### Study design and Setting

We conducted a multi-site, cross-sectional diagnostic accuracy study. Consecutive adolescents were prospectively invited to participate in our study. Participants were recruited from Can Tho and An Giang provinces in the Mekong Delta region of southern Vietnam. Both provinces have high incidence rates of tuberculosis, at over 100 per 100,000. Study recruitment occurred at the Can Tho Lung Hospital (formerly Can Tho Tuberculosis and Lung Diseases Hospital), a tertiary hospital at the edge of Can Tho City, Can Tho; Long Xuyen City Health Centre, a secondary hospital in Long Xuyen, An Giang; as well as decentralised household screening events in more rural areas of the two provinces organised by the NTP. All testing procedures were conducted at the Can Tho Lung Hospital laboratory by trained laboratory technicians.

### Eligibility criteria

Adolescents aged 10 to 19 years who were recommended to undergo investigation for tuberculosis were eligible to participate. Reasons for tuberculosis investigation include (i) living with a contact who has recently received a tuberculosis diagnosis, (ii) presumptive disease based on signs/symptoms, and (iii) chest x-ray abnormalities. Participants must have been capable of providing informed consent and must not have received tuberculosis treatment in the previous 12 months. Concurrent extrapulmonary tuberculosis was not an exclusion criterion, but the participant must have been recommended for pulmonary tuberculosis assessment.

### Procedures

All participant activities occurred at a single study visit. Participants were asked to not eat or drink for 30 minutes before sample collection. Swabs (Copan FLOQswab 520CS01) were first collected by swabbing the back of the tongue for 15 (to a maximum of 30) seconds with a sweeping and rolling motion. Swab heads were snapped off into dry 2 mL cryovial tubes and stored in a cold box (2-8°C) for transport to the laboratory; all TS were tested within 24 hours. Each participant was then coached to provide two sputum samples. Sputum appearance was documented by study staff as ‘mucopurulent’, ‘saliva’, ‘blood-tinged’, or ‘food debris’ and samples were tested regardless of appearance. After providing TS and sputum, participants were asked to complete a short survey of structured questions pertaining to their experience of sample collection.

Separately, laboratory technicians who collected swabs and/or performed TS Xpert for the study completed short semi-structured interviews. Technicians were asked pre-specified questions regarding perceived TS sampling feasibility, processing, and running.

Our index test was TS Xpert Ultra. Briefly, once brought to the laboratory, 700 μL of 2:1 Cepheid sample reagent: phosphate buffer solution was adding to each TS-containing cryotube. TS-buffer tubes were vortexed and then incubated for 15 minutes. Phosphate buffer (1.5 mL) was added to each Xpert cartridge. After TS incubation was completed, 0.5 mL of fluid from TS-buffer tube was pipetted into the buffer-containing cartridge. Xpert tests were then run normally on the GeneXpert platform. At the time of index testing, laboratory staff were blinded to participant’s disease status.

Digital or conventional chest radiography were completed following local protocols for each participant and were read by an experienced physician or tuberculosis doctor. Images were scored as ‘normal’, ‘abnormal, inconsistent with tuberculosis, or ‘abnormal, consistent with tuberculosis.

Each sputum was run on liquid culture (BD BACTEC MGIT, BD, USA) or Xpert Ultra; all samples were tested fresh without freezing. The composite reference standard was defined as either a positive result for ‘MTB detection’ on sputum Xpert, or confirmed growth of *Mycobacterium tuberculosis* (*M. tuberculosis*) on liquid culture. Trace results were considered positive. Reference standard testing was performed according to manufacturer’s instructions. Study data were managed using Research Electronic Data Capture (REDCap) hosted at the University of Sydney Vietnam Institute (10).

### Statistical analyses

The primary outcome measures were sensitivity and specificity with accompanying 90% confidence intervals for TS Xpert. Secondary outcomes included diagnostic yield of TS Xpert; rates of error and invalid results; acceptability of TS sampling by participants; and perceived acceptability of TS Xpert testing by laboratory workers. A precision-based approach was used to estimate sample size (11, 12); to calculate 90% confidence intervals (CI) with a width of ±20% around sensitivity estimate, we estimated we would need 206 individuals. To account for possible unsuccessful test results, we aimed to recruit 225 participants.

Demographic features were summarised using descriptive statistics. Sensitivity and specificity were calculated following the intention-to-diagnose principle (13, 14), with unsuccessful results among reference standard-positive or -negative groups analysed within that disease stratum only (13). A complete case analysis (i.e., individuals with valid positive or negative results on initial test runs only) was also conducted. Diagnostic yield was calculated among all diagnosed with tuberculosis and among all tested (15), and among those only producing salivary sputum. Wilcoxin rank-sum tests were used to test differences between participants’ experiences and acceptability of TS and sputum sampling. Tabular analyses were conducted using ‘epiR’ (16). All analyses were run using R version 4.5.1 (17).

### Ethical issues

Ethical approval was obtained from the University of Sydney Human Research Ethics Committee (2024/HE001890), as well as Institutional Review Board (IRB), Can Tho Lung Hospital (then known as Can Tho Tuberculosis and Lung Diseases Hospital) (483/QÐ – BVLBP).

All participants provided informed consent. Per local regulations, for adolescents aged 10 to 15, participant verbal assent and parental consent were obtained; for participants aged 16 and 17, written participant and parental consent were obtained. Participants found to have tuberculosis on reference standard testing were referred for usual care in the Vietnamese NTP. Index test results did not influence usual care.

## Results

Between 2 July and 26 December 2025, we enrolled 225 adolescents aged 10 to 19 years (Figure 1), of whom 157 (70%) were close contacts of people with newly diagnosed tuberculosis; 63 (28%) presented with presumptive tuberculosis; and 5 (2%) had abnormal chest x-rays. Median (IQR) participant age was 16 (13, 18) (Figure S1) and a minority of participants (96/225, 43%) were symptomatic (Table 1); cough was the most frequently reported symptom (Table S1), with a median duration of 1 week. All participants were investigated for pulmonary tuberculosis without concurrent extrapulmonary tuberculosis. No one reported previously receiving tuberculosis treatment.

**Figure 1:**
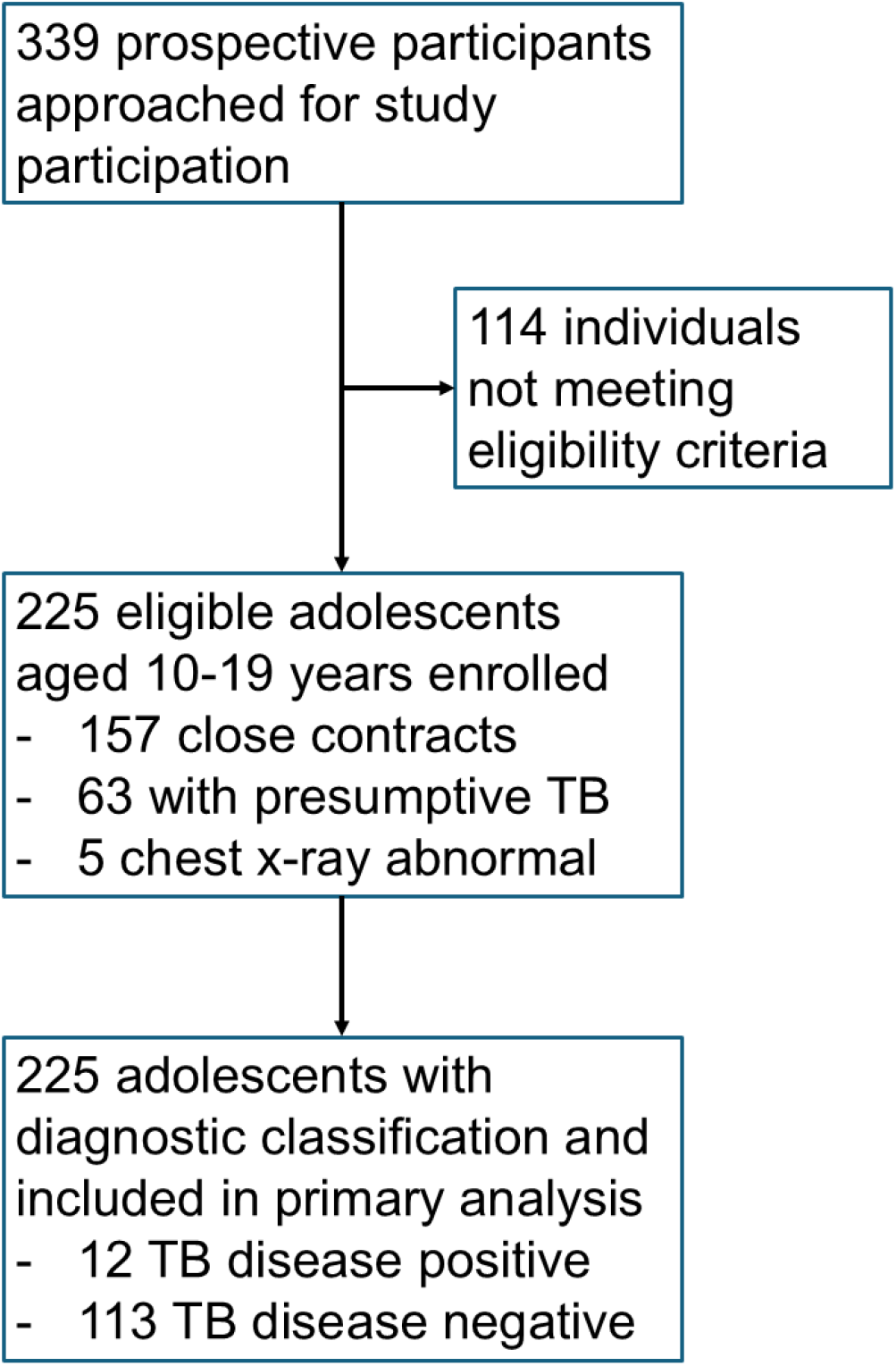
Flow diagram of participants. Individuals were classified as TB disease positive if they had a positive result for TB on liquid culture and/or Xpert MTB/RIF Ultra on sputum samples.

**Table 1:**
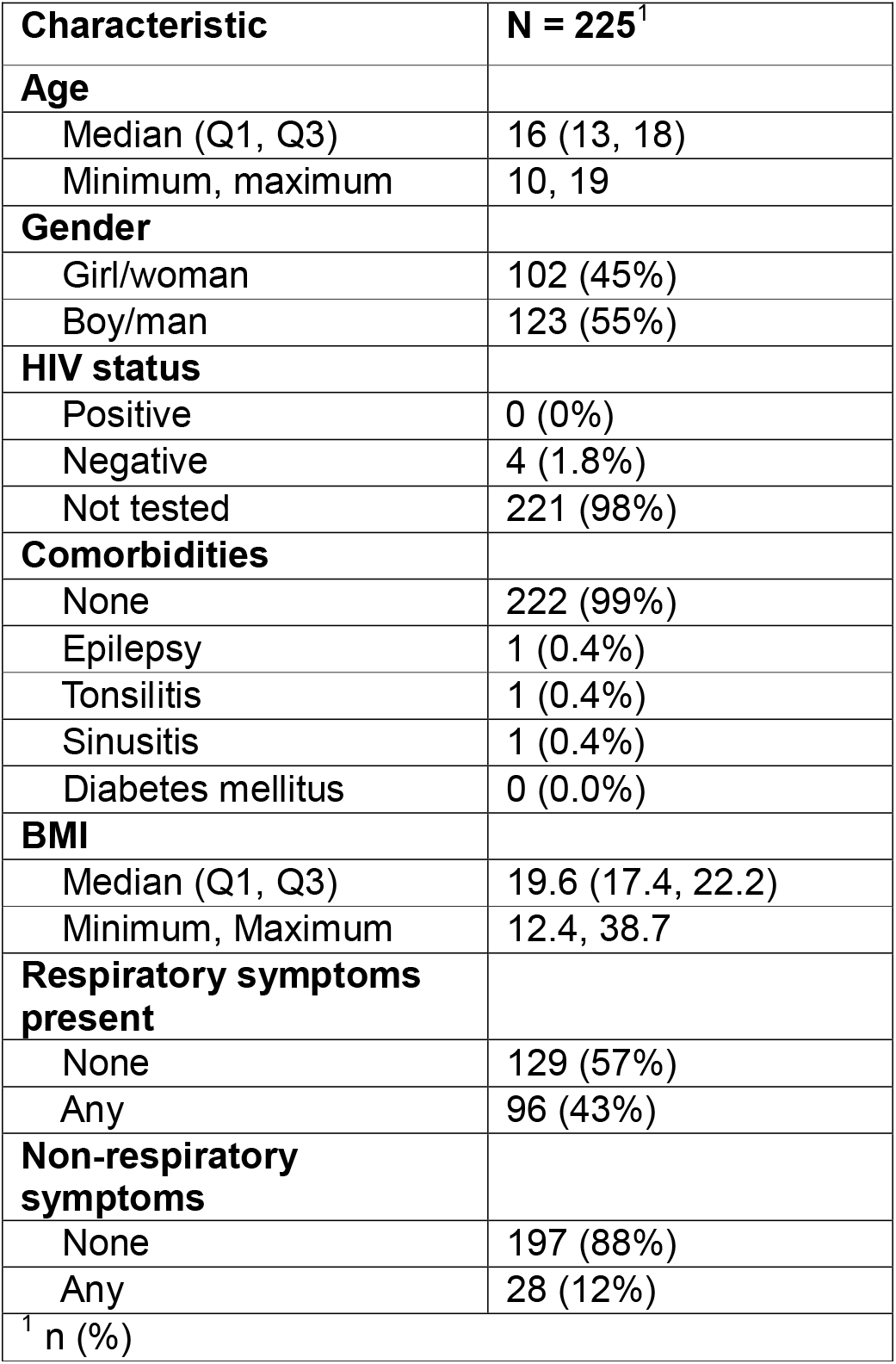
Study population features. Q – quartile.

Tongue swabs were collected from all participants. At least one mucopurulent sputum was collected from 116 participants (51.6%), while 109 could only produce saliva. Younger adolescents aged 10-14 (54/93, 58.1%) were more likely than adolescents aged 15-19 (55/132, 30.3%) to only produce saliva (p=0.0221).

The composite reference standard classified 12 (5.3%) adolescents as tuberculosis-positive, including a trace-positive 13-year-old with chest x-ray abnormalities consistent with tuberculosis. TS Xpert produced one error (0.4%); repeat testing was not possible. Sputum Xpert also produced one error; repeat testing using a second sputum sample yielded a negative result.

Following the intention-to-diagnose principle, TS Xpert sensitivity was 58.3% (90% CI: 35.6, 78.0) while specificity was 99.5% (90% CI: 97.9, 99.9) (Table 2A). Complete case analysis estimated TS Xpert sensitivity of 58.3% (90% CI: 35.6, 78.0) and specificity of 100.0% (90% CI: 98.7, 100) (Table 2B). Among adolescents who could produce a sputum sample, sensitivity increased to 60.0% (90% CI: 35.2, 80.6) and specificity remained 100.0% (90% CI: 97.5, 100.0) (Table 2C), and for those only producing salivary ‘sputum’, sensitivity was 50.0% (90% CI: 12.1, 87.9) with specificity of 99.1% (90% CI: 96.0, 99.8). One participant was found to have rifampicin-resistant tuberculosis on sputum Xpert; TS Xpert did not detect this case of tuberculosis.

**Table 2:** Diagnostic accuracy of tongue swab Xpert Ultra. A: including all participants; B: complete case analysis; C: among participants producing mucopurulent sputum only; D: among participants not producing sputum only.

|  |  |  |  |
| --- | --- | --- | --- |
| A: Intention-to-diagnose analysis<sup>a</sup> (n=225) |  |  |  |
|  |  | Composite reference standard |  |
|  |  | TB positive | TB negative |
| Swab Xpert | Positive | 7 | 0 |
|  | Negative | 5 | 213 |
|  | Unsuccessful* | 0 | 1 |
| B: Complete case analysis<sup>b</sup> (n=222) |  |  |  |
| Swab Xpert | Positive | 7 | 0 |
|  | Negative | 5 | 210 |
|  | Unsuccessful | NA | NA |
| C: Mucopurulent sputum producers (n=116) |  |  |  |
| Swab Xpert | Positive | 6 | 0 |
|  | Negative | 4 | 106 |
|  | Unsuccessful* | 0 | 0 |
| D: Salivary sputum producers (n=109) |  |  |  |
| Swab Xpert | Positive | 1 | 0 |
|  | Negative | 1 | 106 |
|  | Unsuccessful* | 0 | 1 |

<sup>a</sup>: When estimating sensitivity, unsuccessful results among disease-positive are excluded from numerator and included in denominator. When estimating specificity, unsuccessful results among disease-negative are included in numerator and in denominator.
<sup>b</sup>: When estimating sensitivity and specificity, individuals missing a valid result on first run of each test are removed from calculation.
\*For participants requiring repeat testing due to a reference standard component test initially producing an unsuccessful result, the final classification based on at least one valid result was used.

TS Xpert diagnostic yield among all diagnosed with tuberculosis was 58.3% (7/12). Among those only producing salivary sputum, the diagnostic yield among all diagnosed was 50.0% (1/2). The diagnostic yields of TS Xpert and first sputum Xpert among all tested for tuberculosis were 3.1% (7/225) and 4.5% (11/225), respectively, a difference that was not statistically significantly different (p=0.449).

Examining the observed test result patterns (Table 3) shows that TS Xpert results were positive only among participants who had positive sputum Xpert, positive culture, chest x-ray abnormality, and were reportedly symptomatic. Notably, TS Xpert did not detect tuberculosis among two adolescents who were positive on sputum Xpert and culture. TS Xpert was more frequently positive when sputum Xpert semi-quantitative result was high (4/4) or medium (1/2) versus low, very low, or trace (1/4) (Table S2). This indicates that TS Xpert detects more advanced or severe tuberculosis well, but may miss cases earlier in the disease spectrum.

**Table 3:** Frequency of observed test result and symptoms patterns. For participants requiring repeat testing due to a test initially producing an unsuccessful result, the final valid test result was included. C/w: consistent with; TB – tuberculosis; Xpert: Xpert MTB/RIF Ultra; (-): negative result; (+): positive result.

| **Tongue swab Xpert** | **Sputum Xpert** | **Sputum culture** | **Chest x-ray** | **Respiratory symptoms** | **Frequency** |
| --- | --- | --- | --- | --- | --- |
| (-) | (-) | (-) | Normal | None | 118 |
| (-) | (-) | (-) | Normal | Any | 78 |
| (-) | (-) | (-) | Abnormal not c/w TB | None | 6 |
| (-) | (-) | (-) | Abnormal not c/w TB | Any | 6 |
| (-) | (-) | (-) | Abnormal, c/w TB | None | 4 |
| (-) | (-) | (-) | Abnormal, c/w TB | Any | 1 |
| (-) | (-) | (+) | Abnormal, c/w TB | Any | 1 |
| (-) | (+) | (-) | Normal | None | 1 |
| (-) | (+) | (-) | Normal | Any | 1 |
| (-) | (+) | (+) | Abnormal, c/w TB | Any | 2 |
| (+) | (+) | (+) | Abnormal, c/w TB | Any | 7 |

Regarding adolescents’ experiences of providing TS and sputum samples, the time spent to produce a sample, physical comfort level, and complexity of TS collection compared favourably to sputum collection (Table 4). As Xpert is not optimised to run on TS, we also asked whether participants would find it annoying to provide a sample for repeat testing, i.e., if the initial test failed; over 97% strongly disagreed or disagreed that they would find this annoying. TS sampling and sputum were found to be acceptable to most participants, although many more adolescents ‘strongly’ agreed that TS was acceptable (p<0.001).

**Table 4:** Participant experiences with sampling and sample acceptability.

| **Feature** | **Tongue swab**<br>**(n = 225)** | **Sputum**<br>**(n = 225)** | **p-value** |
| --- | --- | --- | --- |
| **Time spent collecting sample was far too long** |  |  |  |
| Strongly disagree | 183 (81%) | 95 (42%) | p < 0.001 |
| Disagree | 33 (15%) | 38 (17%) |  |
| Neutral | 3 (1.3%) | 8 (3.6%) |  |
| Agree | 6 (2.7%) | 69% (31%) |  |
| Strongly agree | 0 (0%) | 15 (6.7%) |  |
| **Sample collection was very physically uncomfortable** |  |  |  |
| Strongly disagree | 203 (90%) | 176 (78%) | p < 0.001 |
| Disagree | 17 (7.6%) | 23 (10%) |  |
| Neutral | 2 (0.9%) | 8 (3.6%) |  |
| Agree | 1 (0.4%) | 15 (6.7%) |  |
| Strongly agree | 2 (0.9%) | 3 (1.3%) |  |
| **Providing sample made me feel very awkward** |  |  |  |
| Strongly disagree | 185 (82%) | 169 (75%) | p < 0.13 |
| Disagree | 20 (8.9%) | 31 (14%) |  |
| Neutral | 4 (1.8%) | 5 (2.2%) |  |
| Agree | 13 (5.8%) | 15 (6.7%) |  |
| Strongly agree | 3 (1.3%) | 5 (2.2%) |  |
| **Sample collection process was really complicated** |  |  |  |
| Strongly disagree | 200 (89%) | 174 (77%) | p < 0.001 |
| Disagree | 18 (8.0%) | 25 (11%) |  |
| Neutral | 4 (1.8%) | 4 (1.8%) |  |
| Agree | 2 (0.9%) | 19 (8.0%) |  |
| Strongly agree | 1 (0.4%) | 4 (1.8%) |  |
| **Repeat sampling would be very annoying to me** |  |  |  |
| Strongly disagree | 187 (83%) | 158 (70%) | p < 0.001 |
| Disagree | 24 (11%) | 28 (12%) |  |
| Neutral | 8 (3.6%) | 11 (4.9%) |  |
| Agree | 4 (1.8%) | 21 (9.3%) |  |
| Strongly agree | 2 (0.9%) | 7 (3.1%) |  |
| **Overall, this sample was acceptable to me** |  |  |  |
| Strongly disagree | 6 (2.7%) | 6 (2.7%) | p < 0.001 |
| Disagree | 4 (1.8%) | 11 (4.9%) |  |
| Neutral | 1 (0.4%) | 10 (4.4%) |  |
| Agree | 53 (24%) | 82 (36%) |  |
| Strongly agree | 161 (72%) | 116 (52%) |  |

We completed semi-structured interviews with three laboratory technicians. Overall, TS collection and testing on Xpert was highly acceptable to technicians. They also perceived TS collection as acceptable to sampled adolescents, who could readily understand the procedure and sit still for its duration. Some adolescents seemed ‘sightly shy’ or perhaps ‘queasy’, but the process was perceived as being easier for participants than obtaining a sputum. As one technician noted: ‘after tongue swab, patients complain about difficulty expectorating sputum when doing sputum next; no complaints about tongue swab.’

There were logistical challenges to untangle regarding transporting and testing samples within 24 hours, particularly when there were high volumes of TS for testing from contact screening events. The technicians suggested that developing a method to preserve TS for over 24 hours would be welcome. Some concern was expressed regarding contamination risk to the swab collector, and technicians felt that the risk of contamination to them during TS collection was higher than when collecting sputum. Using a droplet barrier, standing to the side of the participant while swabbing, wearing PPE, and working in a well-ventilated environment were felt to mitigate, if not obviate, risks.

Preparing the TS sample for Xpert testing was found to be slightly more complex than sputum preparation, and the risk of contamination during TS manipulation was seen as lower than with sputum. One technician suggested that a pre-made buffer at the correct dilution would be welcome and quicker, and suggested setting up clear processes for managing Xpert cartridges of different samples (e.g., using separate trays, batching, etc).

## Discussion

In our consecutively recruited population of adolescents recommended to undergo tuberculosis investigation, TS Xpert had low sensitivity, with excellent specificity. Additionally, TS sampling was found to be highly acceptable to participants and laboratory staff.

Molecular testing for tuberculosis using tongue swabs is a major advance in the fight to end tuberculosis and find the ‘missing millions’, as TS could be collected from, essentially, all individuals, even when respiratory samples are unavailable. However, we observed lower sensitivity with TS Xpert than has been reported previously (18, 19), which may be partially attributable to the sample processing methods. As Xpert was not designed to run with TS, research is ongoing to determine the optimal sample preparation protocol with the lowest limit of detection, and it is possible that a different dilution of reagent-buffer solution may have improved sensitivity (20). Additionally, our study population’s median age was notably lower than that of most other published studies (8, 18, 19), and participants may have had correspondingly lower *M. tuberculosis* bacillary loads.

The low sensitivity of TS Xpert we observed demonstrates the importance of careful evaluation among sub-populations who need better-performing tuberculosis tests. New samples and assays must be rigorously assessed in young people (9) as well as other key groups, e.g., people living with HIV (21), to ensure they will work as hoped. This is because the actual burden of *M. tuberculosis* impacts test positivity (22), so an ‘accessible’ sample alone may be insufficient to improve diagnosis in groups with smear-negative or paucibacillary disease (23). The (understandable) tendency to chase new technological ‘silver bullets’ (24) once available is probably not the ultimate cure for the persistent tuberculosis case detection gap. This trend was previously seen during the COVID-19 pandemic when major stakeholders recommended bi-directional testing for tuberculosis and COVID-19 (25, 26) without evidence; subsequently, prospective studies (27, 28) showed this to be an inefficient approach.

Our findings on patient acceptability are broadly aligned with results of another multi-country survey, primarily among adults (29). There, a majority (61%) of participants indicated a preference for TS over sputum. Although we did not query head-to-head whether our participants preferred one sample over another, a larger share of adolescents ‘strongly agreed’ that TS were acceptable to them than sputum. In a qualitative study interviewing caregivers, healthcare workers, and NTP stakeholder regarding oral swab testing among young children (<5 years), swabs were considered highly acceptable. Swabs were viewed favourably as they were not invasive, painful, or time-consuming to collect (30). Our participants also expressed an appreciation for the simplicity and short time required for TS collection, which, coupled with their high accessibility, may bode well for their deployment in screening. Future focussed work is needed for this test use-case.

Our study had some notable strengths. To our knowledge, this is the first study describing performance of TS tuberculosis testing on any molecular platform specifically in adolescents. Designing our study such that only one participant visit was required permitted excellent completion of the patient experience-acceptability survey. By utilising a composite reference standard comprised of both molecular and bacteriological tests, we can be reasonably confident in participants’ disease classification. Our findings are likely generalisable to other high tuberculosis incidence settings, as Vietnam is classified as a country with a high burden of tuberculosis (31).

There were also limitations. We observed a relatively low event rate, resulting in low precision for sensitivity estimates; we acknowledge that tuberculosis prevalence was low, and that this study is just beginning to fill a knowledge gap. We have reported other salient outcomes, but more diagnostic evaluation in this group is necessary. Additionally, the clinical presentation of our study population was heterogenous, with approximately 50% presenting with symptoms. However, our population does represent adolescents who would be recommended for tuberculosis investigation by most NTPs in countries with high tuberculosis burdens.

In a population of adolescents recommended for tuberculosis investigation, participant experiences were positive and sampling acceptability was high. TS Xpert sensitivity was relatively low and specificity was excellent, with TS Xpert seemingly detecting the most advanced cases of tuberculosis. Among those who could not produce mucopurulent sputum, sensitivity was poor. Further work should be undertaken to improve TS molecular testing in adolescents, including potentially as part of diagnostic algorithms.

## Data Availability

Deidentified participant data that underlie the results reported in this Article, with an accompanying data dictionary, can be made available to investigators whose proposed use of the data has been approved by an independent review committee.

## End materials

## Acknowledgements

The study team is grateful to Claudia M. Denkinger for sharing standard operating procedures for tongue swab processing and testing.

## Funding

This study was funded by a Research Support Grant from the Australian Respiratory Council. ELM was supported by a Canadian Institutes of Health Research Fellowship (472823). GJF was supported by the National Health and Medical Research Council (2007920). GH received financial assistance of the European Union (Grant no. DCI-PANAF/2020/420-028), through the African Research Initiative for Scientific Excellence (ARISE), pilot programme. ARISE is implemented by the African Academy of Sciences with support from the European Commission and the African Union Commission. The funders had no role in study design, data collection, data analysis, data interpretation, or writing of the report.

## Declaration of interests

The authors have no conflicts of interest to declare.

## Author contributions

Conceptualisation: ELM

Data curation: LC, KHM, ELM

Data collection: LC, KHM, NTN

Formal analysis: ELM

Funding: ELM, GJF

Methodology: ELM, TTM, GH, NTN

Project management: TTM, PNY

Supervision and oversight: PNY, TAN, GJF, HTT, NTN

Writing – first draft: ELM

Writing – editing and review: All

## Notes

### Competing Interest Statement

The authors have declared no competing interest.

### Author Declarations

Ethical approval was obtained from the University of Sydney Human Research Ethics Committee (2024/HE001890), as well as Institutional Review Board (IRB), Can Tho Lung Hospital (then known as IRB, Can Tho Tuberculosis and Lung Diseases Hospital) (483/QĐ-BVLBP).

