## Supplementary figures and images for "Tongue swab Xpert MTB/RIF Ultra testing for tuberculosis in adolescents: a cross-sectional study of diagnostic accuracy and acceptability"

### Supplemental figure 1

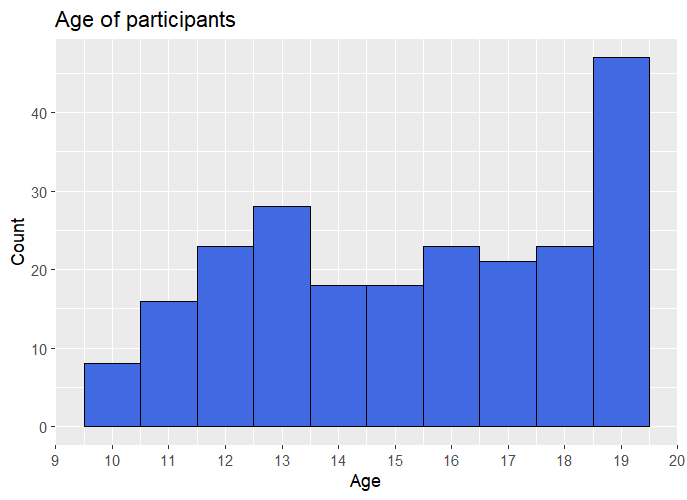
