## Supplemental table 1 for "Tongue swab Xpert MTB/RIF Ultra testing for tuberculosis in adolescents: a cross-sectional study of diagnostic accuracy and acceptability"

**Table S1**: Description of respiratory symptoms reported by enrolled study participants. 96 of 225 participants reported experiencing any respiratory symptom.

| **Respiratory symptom** | **Reported** **N = 96**^1^ |
| --- | --- |
| Cough | 80 (83%) |
| Breathlessness | 7 (7.3%) |
| Haemoptysis | 3 (3.1%) |
| Chest pain | 4 (4.2%) |
| Other respiratory symptoms | 28 (29%) |
| ^1^ n (%) | |
