## Supplemental table 2 for "Tongue swab Xpert MTB/RIF Ultra testing for tuberculosis in adolescents: a cross-sectional study of diagnostic accuracy and acceptability"

**Table S2**: Positive results on tongue swab Xpert Ultra stratified by sputum Xpert Ultra semi-quantitative result. MTB – Mycobacterium tuberculosis; NA – not applicable; Xpert – Xpert MTB/RIF Ultra.

|  |  | **Sputum Xpert semi-quantitative result** | | | | |
| --- | --- | --- | --- | --- | --- | --- |
|  |  | High | Medium | Low | Very low | Trace |
| **Tongue swab Xpert MTB result** | MTB detected | 4 | 2 | 1 | 0 | 0 |
|  | MTB not detected | 0 | 1 | 2 | 0 | 1 |
|  | Error | 0 | 0 | 0 | NA | 0 |
